# Reduced Middle Cerebral Artery Blood Velocity Response during Low-Volume High-Intensity Interval Exercise in Chronic Stroke

**DOI:** 10.1101/2022.12.15.22283530

**Authors:** Alicen A. Whitaker, Saniya Waghmare, Robert N. Montgomery, Stacey E. Aaron, Sarah M. Eickmeyer, Eric D. Vidoni, Sandra A. Billinger

## Abstract

**Background:** High-intensity interval exercise (HIIT) is currently being implemented in stroke rehabilitation to improve walking function. HIIT alternates between high-intensity and recovery bouts, which causes rapid increases and decreases in blood pressure, which may impact cerebrovascular function in people post-stroke. To date, there is no information on the acute cerebrovascular response in people post-stroke. To address this gap in knowledge, we tested whether the cerebrovascular response was reduced when compared to sedentary age- and sex-matched adults (CON) at the following timepoints: 1) baseline (BL), 2) during a single 10-minute bout of low-volume HIIT, 3) immediately following HIIT, and 4) 30 minutes after HIIT.

**Methods:** Middle cerebral artery blood velocity (MCAv), mean arterial pressure (MAP), heart rate (HR), and end tidal carbon dioxide (P_ET_CO_2_) were recorded at all timepoints. A submaximal exercise test was used to determine the estimated maximal workload. The low-volume HIIT bout was performed on a recumbent stepper and consisted of 1-minute high-intensity bouts (70% estimated maximal watts) separated by 1-minute active recovery (10% estimated maximal watts). Statistical analysis used linear mixed models and ANCOVA.

**Results:** Individuals post-stroke demonstrated a reduced MCAv response during HIIT compared to CON (p = 0.03) and MCAv remained lower immediately following HIIT and 30-minutes after HIIT. MCAv decreased below BL immediately following HIIT in both groups (p ≤ 0.04) and returned to BL at 30-minutes after HIIT. No between differences were found for MAP, HR, and P_ET_CO_2_.

**Conclusions:** The key finding from this study suggests a reduced MCAv response during low-volume HIIT in individuals post-stroke compared to age and sex-matched sedentary adults and these differences remained up to 30 minutes following HIIT. Future work is needed to better understand the reduced MCAv response during HIIT despite increased metabolic demand. Clinicaltrials.gov #NCT04673994.

## Introduction

The existing body of literature suggests that high-intensity interval exercise (HIIT) improves walking function post-stroke, although the evidence is not conclusive.^1-3^ HIIT includes bouts of high-intensity exercise followed by active or passive recovery.^4,5^ Our previous work in young healthy adults performing low-volume HIIT with 1-minute intervals showed rapid increases and decreases in heart rate (HR), mean arterial pressure (MAP), and middle cerebral artery blood velocity (MCAv).^6^ Several reviews postulated that HIIT, with rapid fluctuations in MAP may present a unique challenge to the cerebrovascular system in individuals post-stroke, due to the underlying cerebrovascular injury (reviewed in ^7,8^).

Individuals post-stroke demonstrate an impaired steady-state MCAv response during moderate-intensity continuous exercise, potentially due to poor cerebral endothelial function, the presence of vascular disease, poor cerebrovascular reactivity to end tidal carbon dioxide (P_ET_CO_2_) during exercise, presence of stroke or a combination of these factors.^9-12^ Based on our prior work in young healthy adults, we would expect MCAv to increase with exercise onset and then decrease as a result of “arterial acidemia and peripheral chemoreceptor mediated hyperventilation of hypocapnia” typically associated with high-intensity exercise.^6,13^ HIIT may reveal a unique MCAv response in individuals post-stroke due to the repetitive fluctuations in MAP and P_ET_CO_2_.^9,10,12,14^ However, no studies have examined the physiologic response in individuals post-stroke during an acute bout of low-volume HIIT.^14^

To address the gap in knowledge, we characterized the MCAv response to a single bout of low-volume HIIT in individuals with chronic stroke compared to age- and sex-matched controls (CON). We hypothesized that individuals post-stroke would have a reduced MCAv response during HIIT, immediately following HIIT and 30 minutes after HIIT when compared to CON. As an exploratory analysis, we wanted to determine potential participant and stroke characteristics that may be related to greater changes in MCAv or MCAv “responsiveness” during HIIT.

## Methods

Study procedures were approved by the University of Kansas Medical Center Human Subjects Committee. Research was conducted in accordance with the principles embodied in the Declaration of Helsinki and in accordance with local statutory requirements. This study was registered on clinicaltrials.gov (NCT04673994).

We recruited individuals with chronic stroke between 6-months to 5 years and adults without stroke who were matched to sex and age ± 5 years. Inclusion criteria: 1) 40-85 years old, 2) performed < 150 minutes of moderate-intensity exercise per week, and 3) answered consenting questions and followed 2-step command. Exclusion criteria: 1) COVID-19 stroke etiology, 2) symptoms of COVID-19 and/or currently positive, 3) could not stand from a seated position without physical assistance from another person, 4) unable to perform exercise on a recumbent stepper, 5) currently insulin-dependent, 6) currently using supplementary oxygen, and 7) history of another neurologic disorder.

### Study Visit 1

Participants were informed of study procedures, benefits, and risks, and provided voluntary written informed consent. Next, we collected demographics, current medications, modified Rankin scale (mRS),^15^ and the Fugl Meyer Lower Extremity Subscale.^16^ Participants were screened for MCAv signal by an assessor blinded to group.

#### Submaximal Exercise Test

The total body recumbent stepper (TBRS, T5XR NuStep, Inc. Ann Arbor, MI) submaximal exercise test is a valid and reliable measure of fitness.^17-20^ We have previously published on the use of the TBRS submaximal exercise test in stroke^18,21^ and to determine the workload for the acute HIIT exercise bout.^6^ Following the published protocol, participants began exercising at 30 watts and stepped at a constant pace between 95-100 steps per minute (spm). We followed the TBRS submaximal exercise test’s protocol for workload progression.^17-19^ Heart rate (HR, Polar electro Inc, New York) and rating of perceived exertion (RPE) were collected every minute during the submaximal exercise test. Test termination criteria were followed as previously published.^6,17,21^ After the exercise test, participants performed a 2-minute cool-down and then rested for ∼15 minutes to allow HR and blood pressure to return to resting values.

The TBRS submaximal exercise test linear regression equation was then used to calculate the estimated maximal oxygen consumption (estimated□O_2max_).^17^ The linear relationship between workload and HR (i.e. slope) was plotted to determine the estimated maximal watts (estimatedWatt_max_).^20^ For individuals on beta-blockers, predicted maximal HR was calculated from the beta-blocker equation (164 – (0.7 x age)). Similar to previous HIIT protocols in older adults and clinical populations, high-intensity was ∼70% estimatedWatt_max_ and active recovery was 10% estimatedWatt_max_.^5,22-27^

#### Exercise Familiarization

HIIT familiarization included practicing switching every minute between high-intensity (∼70% estimatedWatt_max_) and active recovery (∼10% estimatedWatt_max_), while maintaining a consistent step rate between 95-100 spm, for no more than 10 minutes. Based on previously published HIIT protocols in individuals with chronic stroke, a HR limit of 85% of age predicted maximum HR was used.^28^

### Study Visit 2

All participants were asked to refrain from caffeine for 8 hours,^29^ food for 2 hours,^30^ and vigorous exercise^31^ and alcohol^32^ for 24 hours. The room was dimly lit with a constant temperature (21 - 23°C).^6,13^

#### Carotid Ultrasound

Participants laid supine for 20 minutes while we conducted bilateral Doppler ultrasound scans (GE Healthcare LOGIQ Ultrasound, Chicago, IL) of the common and internal carotid artery. A blinded physician (SE) reviewed images offline and determined the degree of carotid stenosis based on classifications from the Society of Radiologists in Ultrasound.^6,33^

Next, we collected pulse wave velocity (PWV), a measure of peripheral arterial stiffness.^34^ A thigh cuff was placed on the upper right leg and inflated while a sensor was placed on the right carotid pulse. A higher velocity was indicative of greater arterial stiffness.^34^

#### Experimental Procedure

Participants sat quietly for 20-minutes while instrumented with the following equipment: 1) bilateral TCD probes (2-MHz, Multigon Industries Inc, Yonkers, New York) to measure MCAv, 2) a 5-lead electrocardiogram (ECG; Cardiocard, Nasiff Associates, Central Square, New York) to measure HR, 3) a nasal cannula attached to a capnograph (BCI Capnocheck Sleep 9004 Smiths Medical, Dublin, Ohio) to measure P_ET_CO_2_ and respiratory rate (RR), 4) a left middle finger Finometer (Finapres Medical Systems, Amsterdam, the Netherlands) to measure beat-to-beat MAP, and 5) a gold-standard microphoned, brachial automated sphygmomanometer (Tango M2; Suntech, Morrisville, NC) on the right arm to ensure accurate calibration of the Finometer. For participants with left upper extremity spasticity, the Finometer was placed on the right middle finger.

Recordings started with 5-minutes of baseline (BL) seated rest. Participants performed 10-minutes of low-volume HIIT with 1-minute high-intensity (∼70% estimatedWatt_max_) separated by 1-minute active recovery (10% estimatedWatt_max_). As performed previously, we started with 10% estimatedWatt_max_ to avoid a Valsalva maneuver.^6^ Following HIIT, participants performed 2-minute active cool-down with decreased resistance and step rate. After the cool-down, we continued our recording for another 5-minutes of seated rest to measure recovery immediately following HIIT and then 30 minutes after HIIT.

#### Data Acquisition

As previously published, raw data was sampled at 500Hz and collected via an analog-to-digital unit (NI-USB-6212, National Instruments) and custom-written software within MATLAB (v2014a, TheMathworks Inc, Natick, Massachusetts).^6,13^ Our prior work showed no significant difference between right and left MCAv in healthy adults.^13^ Therefore, the left MCAv signal was used for CON. If the left MCA signal was not obtainable, then right MCAv was used.^13^ In individuals post-stroke, we acquired both right and left MCAv.

We calculated 5-minute averages for the variables of interest at BL, immediately following HIIT, and 30 minutes after HIIT. During HIIT, 1-minute averages were calculated for a total of 10 separate time points.^6,35,36^ We calculated pulsatility index (PI; PI = Systolic MCAv – Diastolic MCAv)/ Mean MCAv) as a surrogate measure of cerebrovascular resistance.^12^

#### Power Analysis

Sample size was calculated based on our prior work reporting the MCAv response to moderate-intensity continuous exercise in individuals post-stroke compared to CON, with an effect size of d = 1.36.^10^ We simulated data for 50 participants (1:1 allocation) for every time point with an effect size of 1.36 assumed for each time point. Data sets were simulated 1,000 times and a linear mixed model was fit for every simulated data set. With n = 50 and if our assumptions were correct, we would have 98% power.

#### Data Analysis

Participant characteristics were analyzed using independent t-tests or Mann-Whitney U test for continuous variables, and Fisher’s exact test or Chi-Square test for categorical variables. Assumptions of the models were assessed using residual analyses, e.g., QQ-plots, predicted vs. fitted. BL values were compared between groups using independent t-tests or Mann-Whitney U tests.

To analyze the primary aim, we fit a linear mixed model with MCAv during HIIT as the response, fixed effects for time and quadratic time (minutes) and group (stroke/CON), and a random intercept. We used an autoregressive-1, AR(1), correlation structure and a one-sided test at α = 0.05.^37^ We also fit a linear mixed model for secondary outcomes (MAP, HR, P_ET_CO_2_, and RR) as the response, fixed effects for time and group (stroke/CON), and a random intercept. To determine whether MCAv and other secondary outcomes were decreased during recovery, we used an ANCOVA adjusted for BL. To determine whether outcomes changed from BL during recovery immediately following HIIT and 30-minutes after HIIT, we used paired t-tests or Wilcoxin Signed Rank tests. We also examined PI between groups using Mann Whitney U tests and within groups using Wilcoxin Signed Rank tests.

As an exploratory aim, we calculated the coefficient of variation (CoV = MCAv SD / MCAv average*100) between high-intensity and active recovery to determine MCAv “responsiveness”. We used univariate linear regressions to determine the association of MCAv CoV with age, sex, history of stroke, carotid stenosis, PWV, estimatedVO2max, absolute high-intensity watts, beta-blocker use, statin use, and BL PI. Univariate linear regressions were also used to explore the association of MCAv CoV with stroke specific variables such as months since stroke, ischemic stroke, MCA stroke, mRS score, and Fugl Meyer lower extremity score.

## Results

Sixty individuals were enrolled within the study. Fifty participants completed this study and were included in the primary analysis, shown in **Figure 1**. No adverse events occurred during the study.

**Figure 1.**
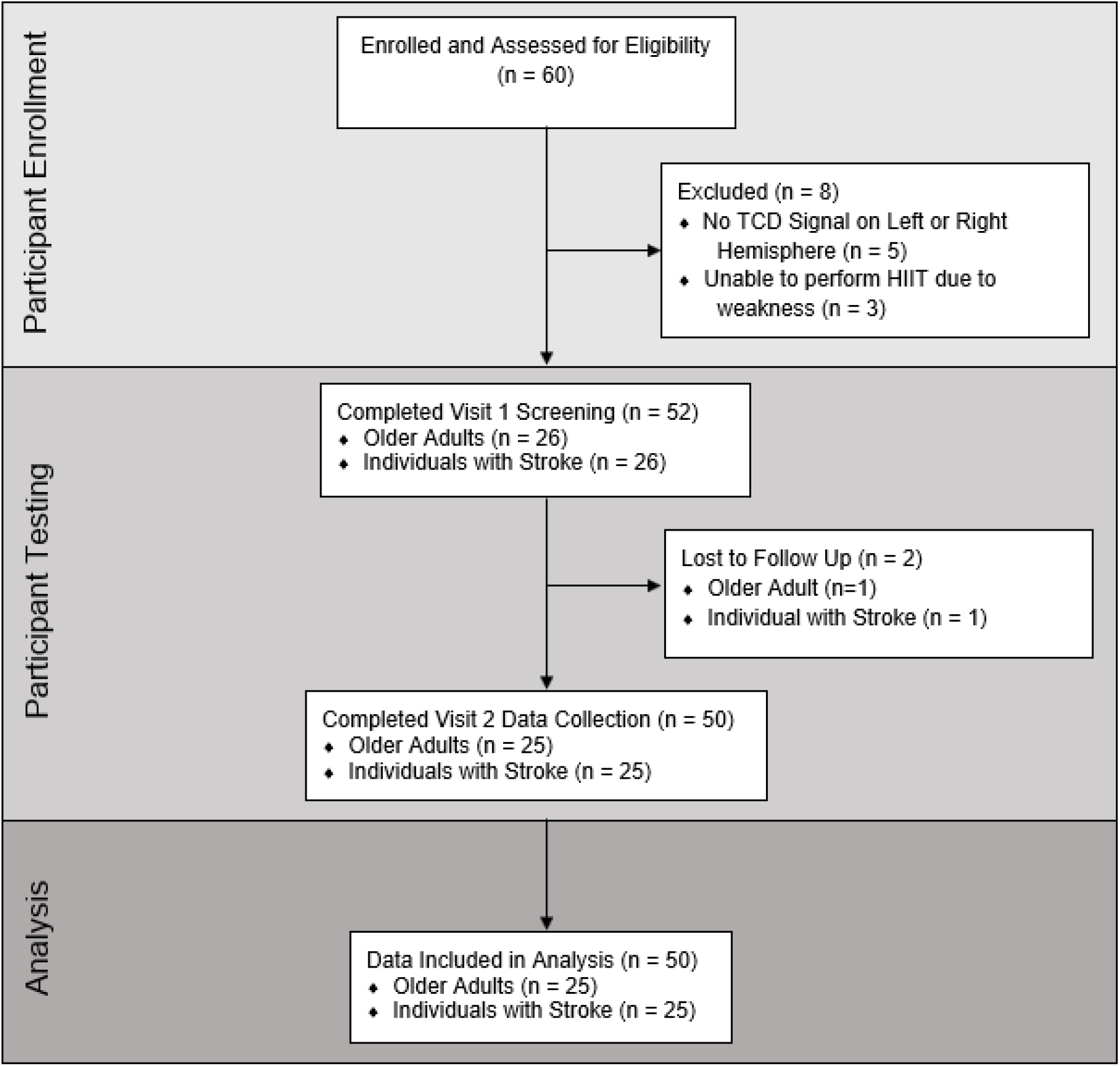
Flow Diagram.

Model assumptions were adequately met using residual analyses. Comparisons of participant demographics between individuals post-stroke (n = 25) and CON (n = 25) are shown in **Table 1**. Participants returned to the laboratory 8 ± 6 days after their first visit. For one individual post-stroke, we were unable to find the ipsilesional MCAv but we were able to find the contralesional MCAv.

**Table 1.**
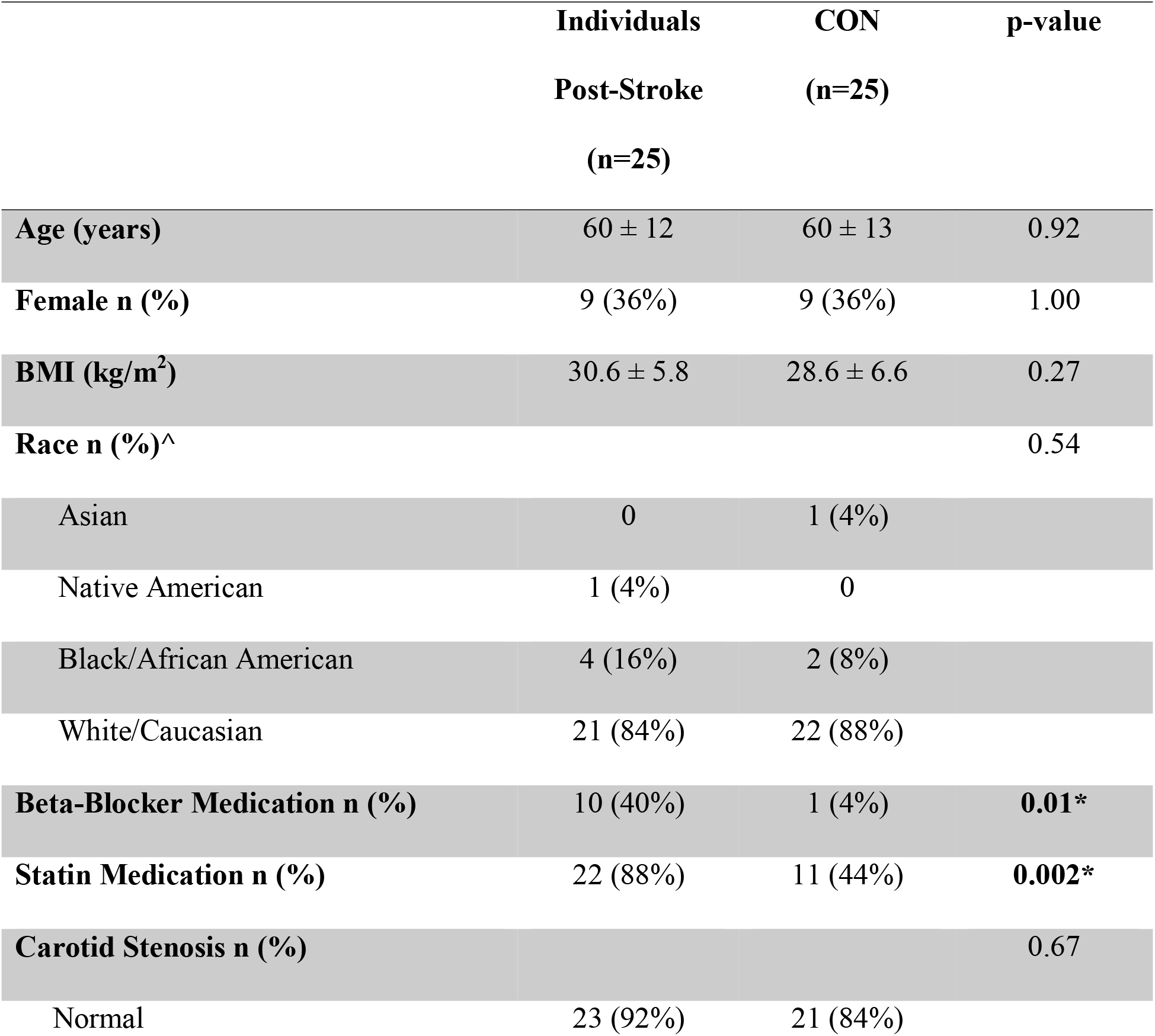
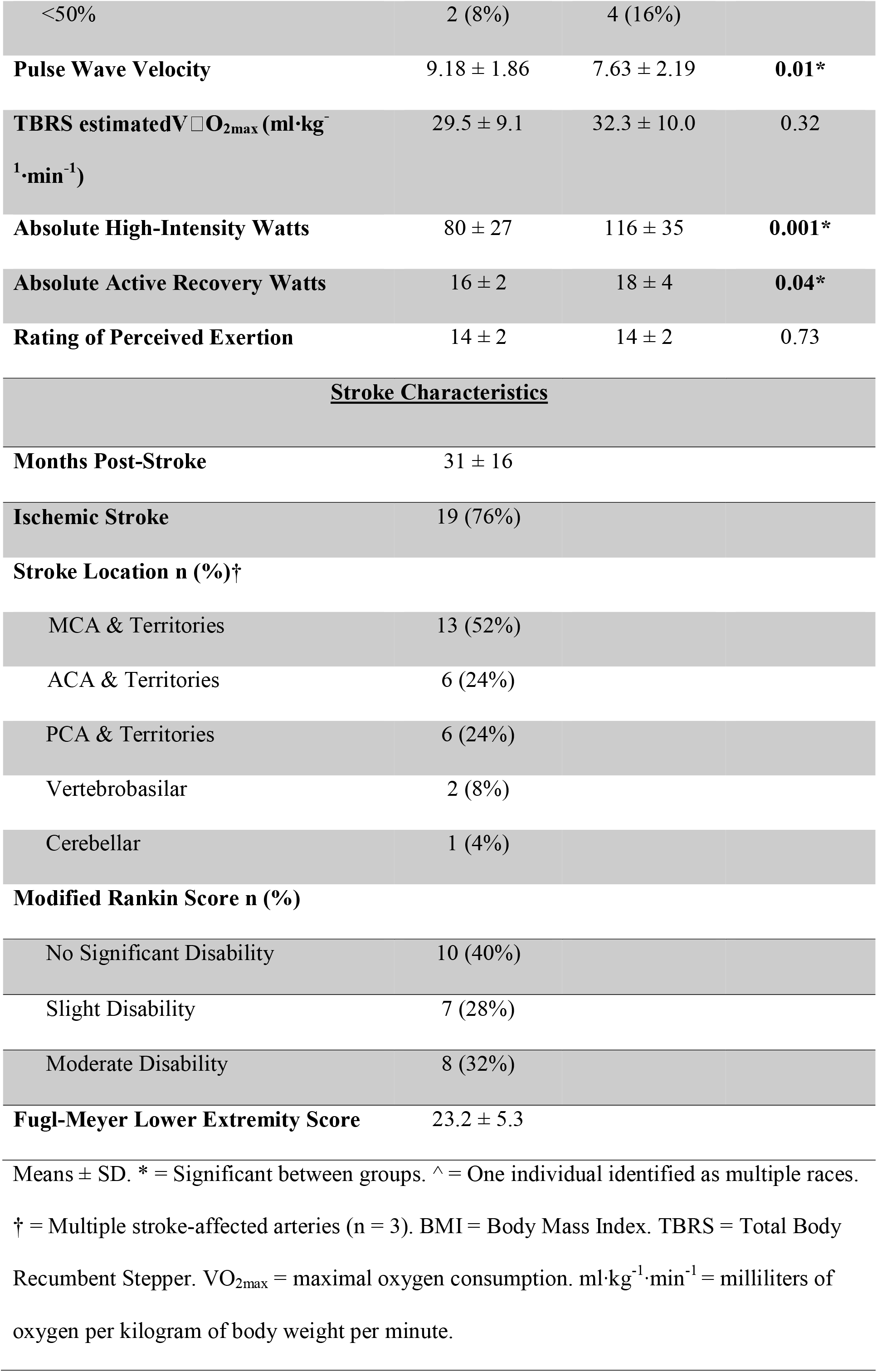
Participant Characteristics.

### MCAv during HIIT and Recovery

At rest, BL MCAv was different between individuals post-stroke 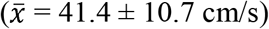 and CON (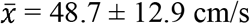, p = 0.04, Cohen’s d = 0.61), shown in **Figure 2**. During HIIT there was evidence for a strong time (p < 0.001) and quadratic time (p < 0.001) effect and a difference between groups 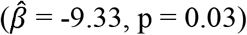. This difference was in the hypothesized direction, with individuals post-stroke having MCAv values 9.33 cm/s lower. As a sensitivity analysis, we included BL MCAv as a covariate in the mixed model. Time and quadratic time were both still associated with MCAv (p < 0.001). However, the group effect was attenuated 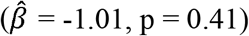. During recovery, MCAv was not different between groups immediately following HIIT (p = 0.42) or 30-minutes after HIIT (p = 0.47), when controlling for BL. In both individuals post-stroke and CON, MCAv decreased lower than BL immediately following HIIT (p ≤ 0.04) and returned to BL at 30-minutes after HIIT (p > 0.14).

**Figure 2.**
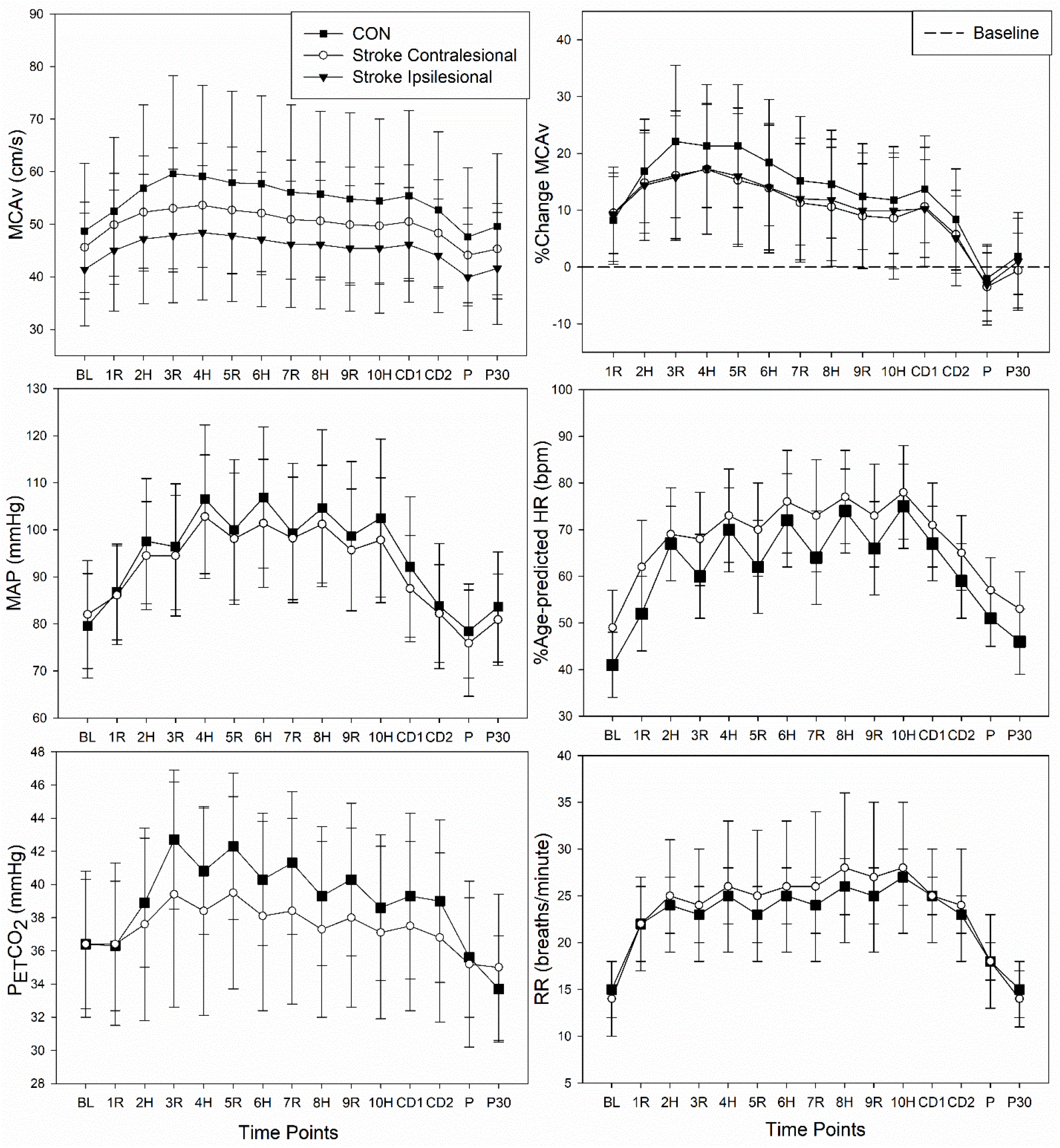
Physiologic Response to HIIT in Individuals Post-Stroke and CON. Controls (CON) = square. Contralesional hemisphere = circle. Ipsilesional hemisphere = triangles. BL = baseline, R = active recovery, H = high-intensity, CD = cool down, P = passive recovery immediately following HIIT, P30 = passive recovery at 30-minutes after HIIT. MCAv = middle cerebral artery blood velocity, MAP = mean arterial pressure, HR = heart rate, P_ET_CO_2_ = expired end-tidal carbon dioxide, RR = respiratory rate.

### Secondary Hemodynamics

No group differences were observed for BL MAP, HR, P_ET_CO_2_, and RR (p > 0.27). During HIIT, all measures increased from BL across time (p < 0.001) and there was no group effect. During recovery immediately following HIIT and 30-minutes after HIIT, there were no group differences in secondary outcomes.

Immediately following HIIT, MAP was lower than BL in individuals post-stroke (p = 0.01), but not CON (p = 0.27). HR was elevated above BL immediately following HIIT in both individuals post-stroke and CON p <0.001). P_ET_CO_2_ was lower than BL immediately following HIIT in individuals post-stroke (p = 0.002) but not CON (p = 0.06). RR also remained elevated above BL immediately following HIIT in both individuals post-stroke and CON p <0.001).

At 30-minutes after HIIT, MAP returned to BL in individuals post-stroke (p = 0.56) but was lower than BL in CON (p = 0.01). HR remained significantly elevated compared to BL in both individuals post-stroke and CON p <0.001). P_ET_CO_2_ was decreased compared to BL in both individuals post-stroke and CON (p <0.001). RR returned to BL values at 30-minutes after HIIT in individuals post-stroke p = 0.99) and CON p = 0.16).

### Pulsatility Index

PI was not different between groups at any timepoint, shown in **Table 2**. Immediately following HIIT, PI was higher than BL in both individuals post stroke and CON (p <0.001). At 30-minutes after HIIT, PI decreased below BL in the CON group (p = 0.001) and returned to BL in individuals post-stroke (p = 0.30).

**Table 2.**
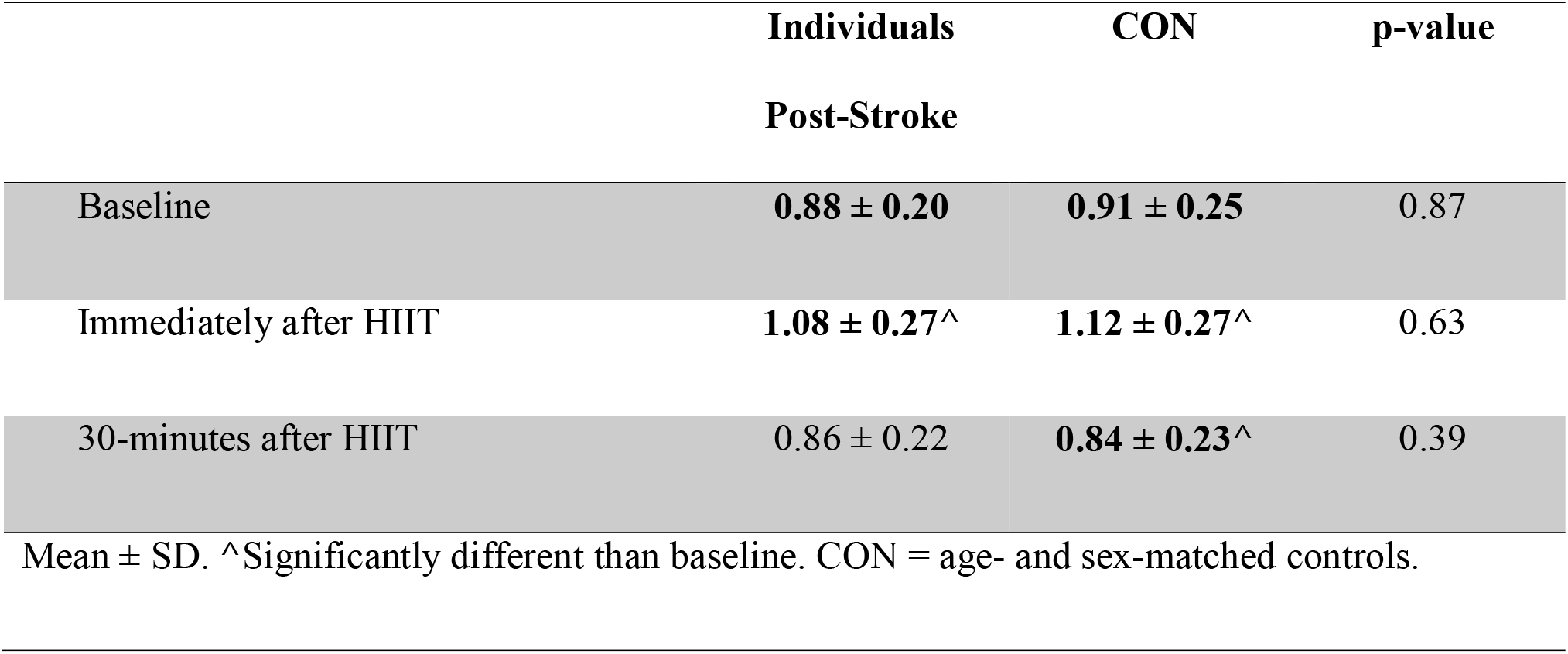
Pulsatility Index.

### MCAv Responsiveness

During HIIT, MCAv CoV was greater in CON 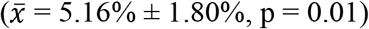 compared to individuals post-stroke 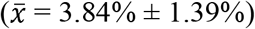. MCAv CoV was associated with sex 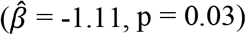, history of stroke 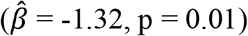, and PWV 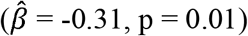, but not age, carotid stenosis, estimatedVO2max, high-intensity Watts, beta-blocker use, statin use, or BL PI. We also found no associations between MCAv CoV and months since stroke, ischemic stroke, MCA stroke, mRS score, and Fugl Meyer lower extremity score.

## Discussion

This is the first study to characterize the MCAv response during an acute bout of HIIT in individuals post-stroke. We report MCAv was uniformly lower throughout HIIT and during recovery compared to age- and sex-matched adults. However, the difference between groups could be accounted for by impaired MCAv at BL. While MCAv was lower at every time point in individuals post-stroke, they showed a pattern comparable to sedentary CON. However, when compared to our previous findings in young healthy adults,^6^ both individuals post-stroke and CON had a blunted MCAv response to HIIT. While age and estimatedVO_2_max were not significantly associated with MCAv responsiveness to HIIT, our prior work in moderate-intensity continuous exercise shows a typical reduction in the MCAv response with age and low physical fitness.^38,39^

To our knowledge, our study was the first to report reduced MCAv at rest in individuals with chronic stroke compared to CON. A reduction in resting MCAv post-stroke compared to CON could be due to stroke pathology or chronic cerebral circulation insufficiency.^40^ Chronic decreases in cerebral blood flow can lead to disruption of glucose use, protein synthesis disorder, and an increased risk of a secondary stroke.^40^ Therefore, we may have identified a potential impairment in resting cerebrovascular health in individuals with chronic stroke.

While not directly measured within this study, cerebrovascular reactivity to carbon dioxide may play an important role in why individuals post-stroke and CON showed blunted MCAv responsiveness during HIIT.^9,41-43^ During HIIT, young healthy adults began to hyperventilate during high-intensity which decreased P_ET_CO_2_ and arterial carbon dioxide (PaCO_2_) causing downstream arteriole vasoconstriction and decreased MCAv.^13^ However, during active recovery, young adults were able to recover their breathing which increased P_ET_CO_2_ and PaCO_2_, causing downstream arteriole vasodilation which “rebounded” or increased MCAv.^6^ While both individuals post-stroke and CON showed a similar P_ET_CO_2_ pattern to young healthy adults during HIIT, MCAv was blunted in both individuals post-stroke and CON.

Some hypothesize that reduced cerebrovascular reactivity with aging and in individuals post-stroke is due to arterial stiffness and poor vasomotor capability.^44^ Our study supports this hypothesis by showing an association between MCAv responsiveness during HIIT and arterial stiffness. Prior studies have also shown that statin use may increase cerebrovascular reactivity to carbon dioxide and the MCAv response to moderate-intensity continuous exercise in individuals post-stroke.^9,11^ However, we did not show an association between statin use and MCAv responsiveness.

MCAv responsiveness was associated with sex, with women having higher MCAv responsiveness. While only 44% of the women within our study were pre-menopausal, our findings are supported by our previous work showing a greater MCAv response to HIIT in young women compared to young men.^6^ Pre-menopausal women have been shown to have greater cerebrovascular reactivity to carbon dioxide compared to men,^45^ which could have contributed to a greater MCAv responsiveness. Reduced MCAv responsiveness in men could also be a protective mechanism, due to having higher resting PI.^46^

Immediately following HIIT, MCAv was decreased below BL which could be due to continued hyperventilation, as shown by decreased P_ET_CO_2_ and increased RR. Breathing and oxygen uptake during exercise recovery is impaired in individuals post-stroke and has been shown to be associated with lower skeletal muscle and arterial stiffness.^47^ When breathing is exacerbated by HIIT, individuals post-stroke may continue to hyperventilate during recovery and decrease P_ET_CO_2_ below BL, causing MCAv to decrease below BL. In individual post-stroke, we found decreased MAP below BL immediately following HIIT. Post-exercise hypotension in individuals post-stroke could challenge the ability of the brain to self-regulate, also known as cerebral autoregulation.^48^

### Pulsatility Index

Immediately following HIIT, PI was elevated in both individuals post-stroke and CON, which we believe may be a typical vascular response, as it follows the pattern of enhanced peripheral arterial tone measured via flow mediated dilation.^49,50^ A previous study in healthy elderly men reported an initial impairment in flow-mediated dilation immediately following HIIT, which suggests an initial vascular challenge.^49^ However, an hour after HIIT, flow-mediated dilation was found to be improved.^49^ It was hypothesized that HIIT may challenge the vascular system initially, but over time HIIT may represent a beneficial stimulus for vascular improvements.^49^ Our study supports this hypothesis with the CON group showing an improved PI at 30-minutes after HIIT compared to BL. However, PI only returned to BL in individuals post-stroke.

### Limitations

While there is controversy as to how much MCA diameter changes with a specific exercise stimulus,^51,52^ TCD is currently the best method to examine the cerebrovascular response to exercise with high temporal resolution.^53^ The middle cerebral artery diameter cannot be measured during exercise and therefore blood velocity was not an exact measure of blood flow. While the TBRS submaximal exercise test allows individuals to perform our HIIT protocol without the need for a maximal exercise test, the prescription of HIIT was based on an estimation rather than directly measuring the anerobic threshold.^4^ We followed previous recommendations of a heart limit of 85% age-predicted maximal HR. Exercising at a higher intensity would have likely decreased P_ET_CO_2_ and MCAv to a greater extent than reported.

### Clinical Relevance

The findings suggest people with chronic stroke are not different in HR, MAP, P_ET_CO_2_, or RR during an acute bout of low-volume HIIT. Our data showed an impaired MCAv response during the HIIT bout in individuals post-stroke when compared to their peers. However, these differences were attenuated when controlling for baseline differences. The blunted cerebrovascular response to HIIT may be due to stroke pathology, poor vascular health, or sedentary behavior.^11^ Future studies are needed to test whether a HIIT intervention could improve cerebrovascular health in individuals post-stroke and sedentary adults. ^54,55^

## Supporting information

STROBE Checklist

COI form

COI Form

COI Form

COI form

COI Form

COI Form

COI Form

## Data Availability

Data are available upon request to the corresponding author.

## Acknowledgements

Thank you, Andrew Geise, Katherine Nguyen, Jake Buchanan, Kailee Carter, Katelyn Struckle, and Emily Hazen for data collection.

